# Closing the Survival Gap: Population-Level Impacts of Digitally-Coordinated Naloxone Distribution on Opioid-Involved Mortality in the Texas Gulf Coast

**DOI:** 10.64898/2026.04.24.26351679

**Authors:** Michael Goodman, Sara Maknojia, Ashley Sciba, David Robertson, Philip Keiser

## Abstract

**Background:** Opioid-related mortality in Texas has escalated dramatically, increasingly driven by illicitly manufactured fentanyl. To address local surges in mortality, the Galveston County Health District deployed the Galveston County Opioid Defense Effort (GCODE) in July 2023, leveraging digitally integrated surveillance data from emergency medical services (EMS) and the Medical Examiner to provide targeted naloxone distribution in identified overdose hot spots.

**Methods:** Using a segmented interrupted time series (ITS) design and Poisson regression with robust standard errors, we evaluated the population-level impact of GCODE on opioid-involved mortality through the end of 2025. Data were sourced from the Galveston Area Ambulance Authority (GAAA) and vital statistics (ICD-10 codes). We assessed mortality trajectory changes, the observed fatality ratio among EMS-detected opioid events (the “Survival Gap”), and demographic and geographic covariates.

**Results:** The Poisson ITS model included 519 weekly observations (N = 14,827 tract-weeks across 101 census tracts). Pre-intervention, opioid mortality increased by 0.16% weekly (IRR = 1.0016; 95% CI: 1.000–1.003; p = 0.011). Following GCODE deployment, the mortality trajectory reversed to a sustained 0.55% weekly decrease (IRR = 0.9945; 95% CI: 0.990–0.999; p = 0.021). The observed fatality ratio among EMS-detected events declined from 7.59% (pre-intervention mean; SD = 0.111) to 1.71% (post-intervention; SD = 0.042; χ² = 19.824; p = 0.0001). Opioid decedents were significantly younger than the general mortality population (OR = 0.945 per year of age; p < 0.001), and were descriptively more likely to lack documented race/ethnicity data (41.23% vs. 8.27% “Unknown”; p < 0.001), limiting equity analysis.

**Conclusions:** The findings are consistent with GCODE having meaningfully reduced opioid mortality by substantially lowering event-level lethality. These results suggest that targeted, digitally coordinated harm reduction can decouple overdose incidence from fatal outcomes, with implications for harm reduction program design in structurally constrained environments.

## Introduction

The United States opioid epidemic has claimed over 600,000 lives over the past two decades, with mortality accelerating sharply due to the proliferation of illicitly manufactured fentanyl and other synthetic opioids (Kiang, Basu, Chen, & Alexander, 2019; Fujita-Imazu et al., 2023). In Texas, opioid-related deaths among adults aged 15–64 surged by 402% between 1999 and 2019 (Salazar & Huang, 2021). The role of fentanyl has grown dramatically in the intervening years: by 2023, fentanyl accounted for 79.0% of all opioid poisoning deaths and 45.3% of all unintentional drug poisoning deaths statewide, rising from just 17.2% and 8.3% respectively in 2018 (Texas Department of State Health Services [DSHS], 2025). Texas Health Region 6/5S (Southeast Texas, the region containing Galveston County) recorded 526 fentanyl-involved deaths in 2023, the second-highest regional burden in the state and representing 31.5% of all Texas fentanyl deaths that year (DSHS, 2025).

Responding to this crisis is complicated by a hostile structural and policy environment. Texas has the lowest density of opioid use disorder (OUD) treatment programs nationally, with only 1.4 per 100,000 residents (Langabeer et al., 2019). While standing orders permitting naloxone dispensing without an individual prescription exist, pharmacy-level accessibility remains low due to cost barriers and dispensing reluctance, and syringe service programs are legally prohibited statewide (Evoy et al., 2018; Claborn et al., 2023a). These structural constraints severely limit the reach of conventional harm reduction approaches.

The epidemiological literature establishes that high-intensity, geographically targeted naloxone distribution can produce a “decoupling” effect-wherein non-fatal overdoses remain stable while opioid-involved mortality declines (Walley et al., 2013). However, evidence suggests this effect is substantially weakened in the current fentanyl era without highly targeted, data-driven deployment strategies (Doleac & Mukherjee, 2018; Irvine et al., 2019). Broad, untargeted naloxone availability is increasingly insufficient; interventions must actively saturate high-risk settings with trained bystanders and accessible reversal agents.

In response, the Galveston County Health District adapted the digital health tracking infrastructure and public health nursing capacity developed during the COVID-19 pandemic to establish the Galveston County Opioid Defense Effort (GCODE). Fully deployed in July 2023, GCODE is a multi-sector collaboration integrating public health agencies, community-based organizations, emergency medical services, and academic institutions. Its core mechanism relies on real-time digital surveillance-aggregating data from the GAAA and the Medical Examiner-to dynamically identify overdose hot spots and systematically equip qualifying organizations operating in those areas to dispense and track naloxone utilization.

This study evaluates the population-level effectiveness of GCODE through the end of 2025 using a segmented interrupted time series (ITS) design. We assess changes in opioid mortality trajectories, the observed fatality ratio among EMS-detected opioid events (the “Survival Gap”), and the demographic and geographic distribution of overdose risk. Findings are interpreted as consistent with, rather than definitively caused by, the GCODE intervention, given the observational nature of the ITS design.

## Methods

### Study Design and Setting

We employed a segmented interrupted time series (ITS) design to evaluate the population-level impact of GCODE on opioid-involved mortality in Galveston County, Texas. The ITS design is widely considered the strongest quasi-experimental approach for evaluating public health interventions when randomization is not feasible, as it formally controls for pre-existing secular trends and temporally attributes outcome changes to the intervention (Bernal et al., 2016; Spittal et al., 2024). The observation window extended from 2016 through the end of 2025, with the intervention point defined as GCODE’s full operational deployment in week 26 of 2023 (July 2023). The study was conducted in Galveston County, Texas Health Region 6/5s. This analysis did not include a concurrent control county, as no geographically proximate county possessed equivalent EMS-to-mortality data linkage infrastructure; this absence is acknowledged as a limitation and is discussed below.

### The GCODE Intervention

In early 2023, the Galveston County Health District repurposed its COVID-19 digital surveillance tools and community nursing infrastructure to coordinate a localized opioid harm reduction response. The resulting GCODE program operates as a multi-sector collaboration among public health officials, community-based organizations, and academic institutions. The program’s core mechanism involves digital aggregation of EMS dispatch data from the GAAA and death investigation records from the Medical Examiner to produce dynamic geographic hot spot maps of opioid-related overdose activity. Hot spots were operationally defined as census tracts in the top quintile of combined opioid EMS call density and opioid mortality rate over a rolling 12-week surveillance window. Organizations qualifying under established criteria and operating within identified hot spots receive Narcan (naloxone) training and supply, and are integrated into a centralized dispensation and utilization tracking system. Figure 4 illustrates the digital coordination pathway between data sources, the GCODE surveillance platform, community partner organizations, and naloxone dispensation.

### Data Sources and Measures

Outcome data were integrated from multiple local surveillance systems. Opioid-involved mortality was sourced from vital statistics using ICD-10 codes identifying confirmed opioid-related deaths. Overall opioid-related overdose incidence was captured using dispatch and emergency medical services response data from the GAAA. Both series were aggregated to the weekly level and linked to census tract population denominators from the American Community Survey.

The primary mechanistic outcome evaluated was the “Survival Gap,” operationalized as the observed fatality ratio among EMS-detected opioid events (weekly opioid deaths divided by weekly opioid-related EMS calls). This metric directly measures the probability of death per EMS-detected overdose event. Readers should note that this ratio measures lethality conditional on EMS detection and does not capture the full population of overdose events, as successful layperson naloxone reversals may not generate an EMS call. EMS calls serve as a validated proxy for population-level overdose incidence; national surveillance data demonstrate a 0.98 aggregate-level correlation between EMS-observed overdose events and concurrent vital statistics mortality (Friedman et al., 2021). Absolute annual counts of opioid deaths and EMS calls by period are reported in Table 4.

Demographic covariates-including age and race/ethnicity-were extracted from vital statistics records. Geographic risk was characterized using census tract boundaries and Social Vulnerability Index (SVI) metrics from the Centers for Disease Control and Prevention.

### Statistical Analysis

To appropriately model the non-negative integer distribution of weekly opioid death counts, we utilized a segmented Poisson regression model. The formal model equation is:

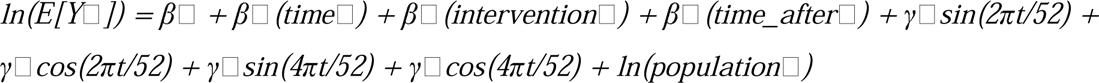

where Y□ is the count of opioid-involved deaths at week t; time□ is a continuous week index (centered at the intervention); intervention□ is a binary indicator (0 = pre-GCODE, 1 = post-GCODE); time_after□ is weeks elapsed since intervention (0 in the pre-period); and the sine/cosine pairs represent two Fourier harmonic pairs (K = 2) capturing annual and semi-annual seasonal variation in overdose incidence. Population at risk (mean census tract population) was included as an exposure offset.

Robust (Huber-White) standard errors were applied to account for potential heteroskedasticity. To correct for residual serial autocorrelation identified by visual inspection of autocorrelation (ACF) and partial autocorrelation (PACF) function plots and confirmed by Breusch-Godfrey and Ljung-Box Q tests, Newey-West robust standard errors were applied with a lag bandwidth of 4 weeks, corresponding to the minimum autocorrelation lag identified in the pre-intervention ACF. The overall model fit was assessed by Wald χ². As a sensitivity analysis, negative binomial regression was estimated to assess overdispersion; the likelihood ratio test of alpha = 0 was non-significant (p = 0.500; lnalpha = −13.19; alpha ≈ 1.87×10□□), confirming the Poisson specification was appropriate.

The model was specified a priori to distinguish: (1) an immediate level change-estimating the abrupt step-change in mortality at the July 2023 intervention point; and (2) a slope change-estimating the gradual, sustained alteration in mortality trajectory following GCODE deployment (Bernal et al., 2016).

For the mixed-effects model examining demographic predictors of opioid-involved death across census tracts, a multilevel logistic regression was employed (melogit in Stata 17), with census tract as the grouping variable (N = 37,372 decedents across 102 tracts; Wald χ²(4) = 1367.82; p < 0.0001). Results are reported as odds ratios (OR) with 95% confidence intervals.

Nonparametric Kruskal-Wallis tests were used as a supplementary unadjusted descriptive comparison for the observed fatality ratio pre/post, but not for primary inference; this approach is acknowledged to be insensitive to temporal ordering, secular trends, and serial autocorrelation (Spittal et al., 2024). All primary inferences derive from the parameterized segmented Poisson model. All analyses were conducted in Stata 17.

## Results

### ITS Analysis of Opioid Mortality Trends

The segmented Poisson ITS model was estimated on 519 weekly observations spanning 2016–2025 (N = 14,827 census tract-week observations across 101 tracts; Wald χ²(4) = 45.54; p < 0.0001). Figure 1 displays the observed monthly opioid death rate across the full study period, with the July 2023 intervention marked. Figure 2 presents the modeled pre- and post-intervention trend lines with fitted values.

**Figure 1.**
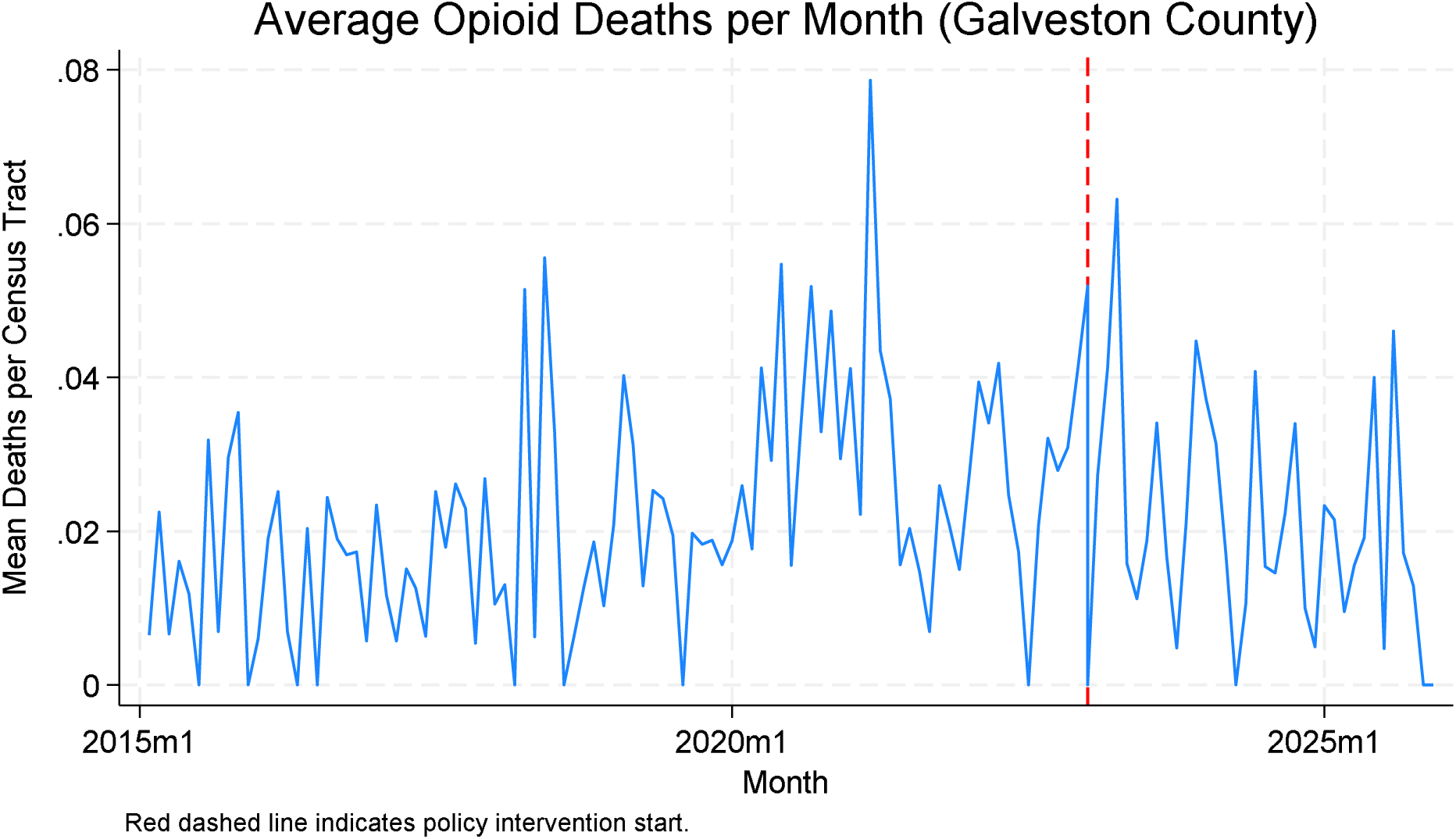
Average monthly opioid death rate in Galveston County, 2015–2025. The observation window extends through end of 2025; the x-axis label "2026m1" reflects weekly data plotted on a monthly scale. Red dashed line indicates GCODE deployment (July 2023).

**Figure 2.**
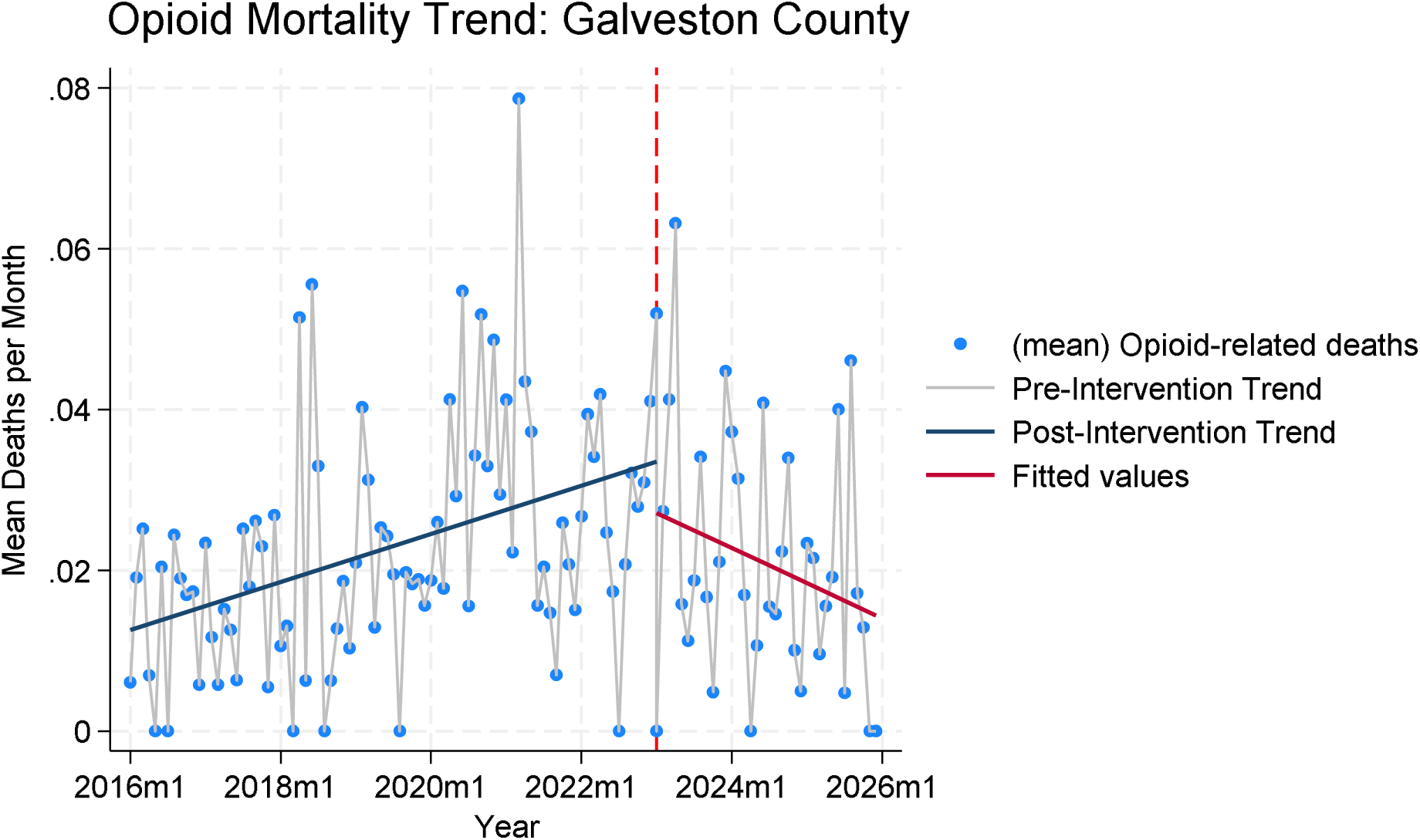
Interrupted time series model of opioid mortality in Galveston County. Blue scatter points represent observed mean opioid-related deaths per week. Gray line shows the pre-intervention modeled trend; dark blue line shows the post-intervention modeled trend. Red line indicates model fitted values. Dashed vertical red line marks GCODE deployment (July 2023). The x-axis extends through 2026 reflecting the final weeks of the 2025 observation year.

During the pre-intervention period (2016–2023w25), opioid-involved mortality exhibited a statistically significant baseline upward trend, increasing at a rate of 0.16% per week (IRR = 1.0016; 95% CI: 1.000–1.003; p = 0.011). In absolute terms, the pre-intervention period recorded a mean of approximately 2.1 opioid deaths per month (see Table 4). At the point of GCODE’s full deployment, the immediate level shift in mortality was not statistically significant (IRR = 0.867; 95% CI: 0.557–1.351; p = 0.529), indicating no abrupt step-change at the intervention moment. However, the intervention was associated with a significant sustained post-rollout slope change (IRR = 0.9945; 95% CI: 0.990–0.999; p = 0.021), representing a reversal to a sustained 0.55% weekly reduction in opioid-involved deaths. The negative binomial sensitivity model yielded substantively identical estimates, confirming that overdispersion did not meaningfully affect inference. Full model estimates are presented in Table 1.

**Table 1.** Interrupted Time Series Analysis of Opioid-Involved Mortality - Segmented Poisson Regression (Primary) and Negative Binomial Sensitivity Model.

| Parameter | IRR (Poisson) | 95% CI | p | IRR (Neg. Binomial) |
| --- | --- | --- | --- | --- |
| Baseline Weekly Trend (Pre-Intervention) | 1.0016 | [1.000, 1.003] | 0.011 | 1.0014 [1.000, 1.003];<br>$p = 0.016$ |
| Immediate Level Shift at Intervention | 0.867 | [0.557, 1.351] | 0.529 | Not significant |
| Post-Intervention Slope Change (Weekly) | 0.9945 | [0.990, 0.999] | 0.021 | 0.9945 [0.990, 0.999];<br>p = 0.019 |
| Overall Model Fit (Wald $\chi^2$) | 45.54 | - | <0.0001 | - |

IRR = incidence rate ratio. Model: 519 weekly observations, population exposure offset, robust standard errors, Newey-West correction (lag bandwidth = 4 weeks), 2 Fourier harmonic pairs for seasonal adjustment. Negative binomial LR test alpha = 0: p = 0.500, confirming Poisson specification appropriate.

### Mechanistic Findings: The Survival Gap

To assess the intervention’s impact on event-level lethality, we analyzed the observed fatality ratio among EMS-detected opioid events (the “Survival Gap”). Figure 3 displays this ratio across the pre- and post-rollout periods. The mean observed fatality ratio declined from a pre-intervention baseline of 7.59% (210 weeks; SD = 0.111) to a post-intervention rate of 1.71% (123 weeks; SD = 0.042). A Kruskal-Wallis equality-of-populations test, used as a supplementary unadjusted comparison, confirmed this reduction was statistically significant (χ² = 19.824; p = 0.0001), representing a 77.5% relative reduction in the observed death risk per EMS-detected event.

**Figure 3.**
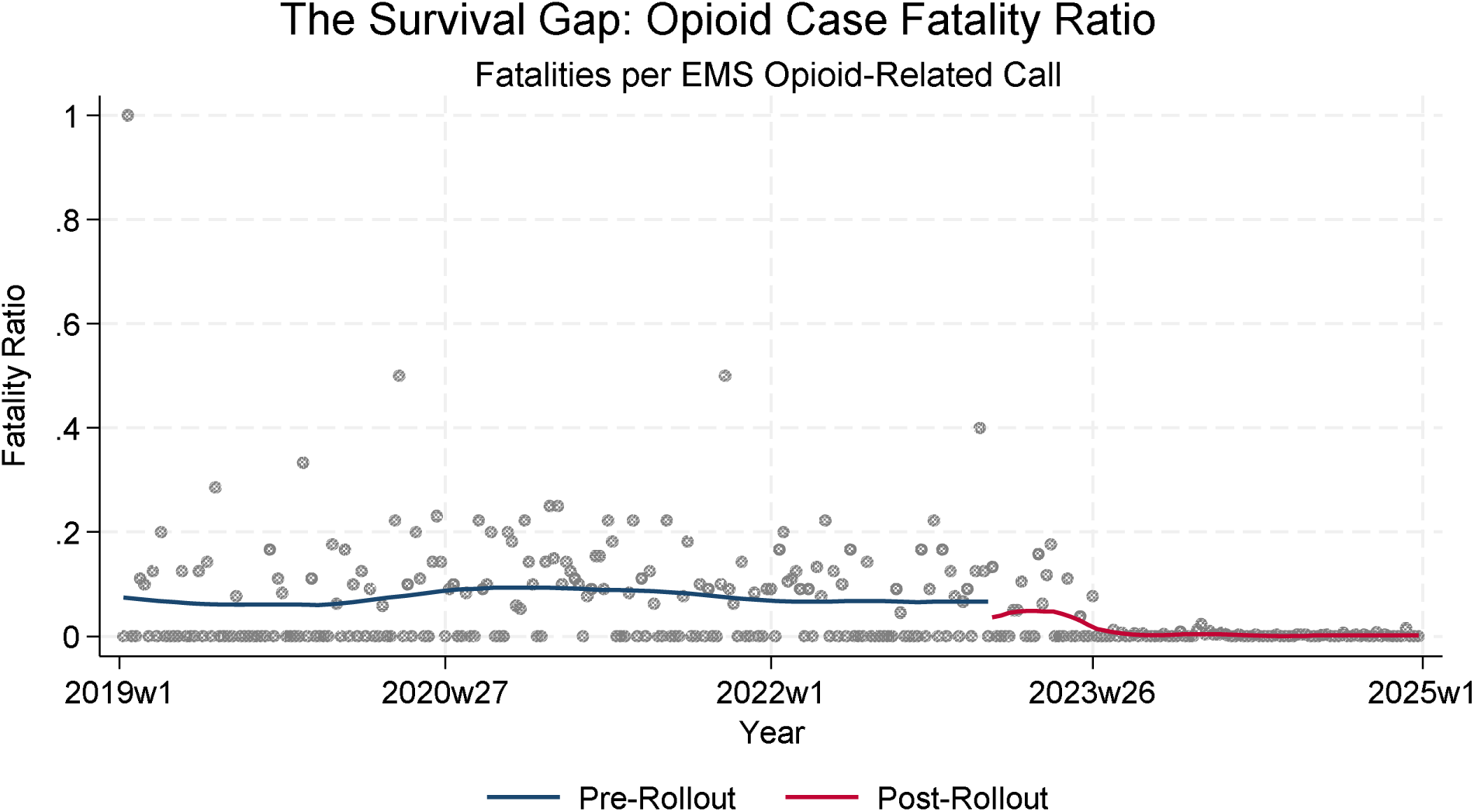
Observed fatality ratio among EMS-detected opioid events (Survival Gap), Galveston County, 2019–2025. Blue line: smoothed pre-rollout trend. Red line: post-rollout trend. Gray scatter points: observed weekly ratios. This metric reflects lethality among EMS-detected events; it does not capture the full population of overdose events including layperson-managed reversals not resulting in an EMS call.

Table 4 presents the absolute counts of opioid deaths and EMS calls in each period, allowing independent assessment of numerator and denominator trends. Readers should interpret the Survival Gap in light of the possibility that increased community naloxone availability may have concurrently reduced EMS utilization through successful layperson reversals, which would tend to reduce the denominator and may partially account for the observed ratio decline.

### Demographic and Geographic Findings

Mixed-effects logistic regression (melogit) modeling decedent characteristics across 102 census tracts (N = 37,372 decedents; Wald χ²(4) = 1,367.82; p < 0.0001) identified a strong and consistent age effect: opioid-involved deaths were descriptively concentrated among younger decedents, with each additional year of age associated with reduced odds of an opioid-involved compared to non-opioid death (OR = 0.945 per year; 95% CI: 0.941–0.948; p < 0.001). While census-tract level random effects were statistically significant (LR test vs. logistic: χ² = 6.86; p = 0.0044), geographic and SVI factors were secondary to age in the overall model. These demographic findings are presented as descriptive rather than inferential, given the data limitations described below.

The racial/ethnic composition of all decedents within the analysis window (2019–2025; N = 15,949) was as follows: White 71.6% (n = 11,426), Black 14.1% (n = 2,251), Other/Unknown 13.6% (n = 2,174), Native American/Alaska Native/Pacific Islander 0.5% (n = 84), and Asian 0.1% (n = 14). The melogit model used White as the reference category (racial identity code 1). The “Other” category (code 4; 13.6%) carried a markedly elevated OR of 6.88 (95% CI: 5.76–8.22; p < 0.001) for opioid-involved versus non-opioid death, substantially exceeding the reference category. This “Other” group likely captures a heterogeneous mix of individuals including those with incomplete racial identification in vital statistics, and its elevated OR is most plausibly interpreted as a marker of demographic data quality failure rather than a genuine racial risk differential. Black decedents showed a significantly lower OR compared to White decedents (OR = 0.633; 95% CI: 0.475–0.844; p = 0.002) within the recorded sample, though this estimate must be interpreted cautiously given the pervasive missingness. All race/ethnicity findings are descriptive and should not be used to draw inferential conclusions about differential population risk. Table 3 presents these findings.

**Table 2.** Pre/Post Comparison of the Observed Fatality Ratio Among EMS-Detected Opioid Events (Survival Gap)

| Metric | Pre-Rollout | Post-Rollout | p-value |
| --- | --- | --- | --- |
| Weeks Observed | 210 | 123 | - |
| Mean Fatality Ratio (deaths / EMS call) | 7.59% | 1.71% | 0.0001 |
| Standard Deviation | 0.111 | 0.042 | - |
| Kruskal-Wallis $\chi^2$ | - | 19.824 | 0.0001 |
| Relative Reduction in Observed Death Risk | - | 77.5% | - |

Analysis window: 2019w1–2025w23. Kruskal-Wallis test used for supplementary unadjusted comparison only; primary causal inference from the segmented Poisson ITS model. Observed fatality ratio = weekly opioid deaths / weekly opioid-related EMS calls (EMS-detected events only).

**Table 3.** Decedent Demographic Characteristics: All Deaths in Analysis Window, Galveston County (Descriptive)

| Characteristic | N (% of all decedents) | OR<sup>□</sup> for Opioid vs. Non-Opioid Death | p-value |
| --- | --- | --- | --- |
| **Total decedents in analysis window** | 15,949 | - | - |
| **Race/Ethnicity<sup>□</sup> (descriptive)** |  |  |  |
| White (reference) | 11,426 (71.6%) | 1.00 (ref) | - |
| Black | 2,251 (14.1%) | 0.633 [0.475–0.844] | 0.002 |
| Other / Unknown<sup>□</sup> | 2,174 (13.6%) | 6.882 [5.762–8.222] | <0.001 |
| Native American / Alaska Native / Pacific Islander | 84 (0.5%) | 0.350 [0.047–2.599] | 0.305 |
| Asian | 14 (0.1%) | Omitted<sup>□</sup> | - |
| Age: OR per additional year of age<sup>□</sup> | - | 0.945 [0.941–0.948] | <0.001 |

<sup>□</sup> Mixed-effects logistic regression (melogit); N = 37,372 across full study period; 102 census tracts; Wald $\chi^2(4) = 1,367.82$ . <sup>□</sup> Descriptive proportions from analysis window (2019–2025; N = 15,949). <sup>□</sup> “Other/Unknown” elevated OR most likely reflects demographic data missingness and coding heterogeneity in vital statistics rather than a genuine independent risk factor; interpret with caution. <sup>□</sup> Asian category (n = 14) omitted from regression due to perfect prediction of non-opioid outcome. <sup>□</sup> Each additional year of age associated with 5.5% reduced odds of opioid vs. non-opioid death.

## Discussion

The deployment of GCODE in Galveston County was associated with a meaningful reversal in opioid-involved mortality, with the segmented Poisson ITS model showing a shift from a baseline weekly increase of 0.16% to a sustained post-intervention reduction of 0.55% per week. While the observational ITS design precludes definitive causal attribution and alternative explanations cannot be definitively excluded, the temporal specificity of the slope change-closely aligned with GCODE’s deployment-is consistent with a genuine population-level intervention effect (Bernal et al., 2016; Spittal et al., 2024).

The primary mechanistic pathway consistent with this mortality reduction is illuminated by the Survival Gap. The 77.5% relative reduction in the observed fatality ratio among EMS-detected opioid events-from 7.59% to 1.71%-is consistent with the “decoupling” of fatal and non-fatal overdoses: a phenomenon wherein the proportion of overdose events resulting in death declines as community naloxone availability increases (Walley et al., 2013). This aligns with the overdose mortality decomposition framework, which models population overdose deaths as a multiplicative function of the at-risk population size, overdose frequency, and the probability of death per event (Skinner et al., 2024). The observed reduction in the fatality ratio is potentially mediated through increased community naloxone access and bystander activation in high-risk settings, though contributions from other concurrent changes in the local risk environment-such as shifts in the fentanyl-to-adulterant composition of the local drug supply-cannot be ruled out.

Readers should interpret the Survival Gap metric with appropriate caution. Because the numerator (opioid deaths) and denominator (EMS calls) are both drawn from the same regional context, they are subject to the same potential regional shocks-including changes in local drug supply toxicity or shifts in law enforcement activity-and therefore do not constitute fully independent streams of evidence. The ITS mortality model and the Survival Gap analysis are best understood as providing consistent directional evidence rather than independent verification. Table 4 presents absolute counts of deaths and EMS calls by period to support evaluation of underlying trends in each series separately.

**Table 4.** Absolute Counts of Opioid-Involved Deaths and EMS Calls by Study Period□.

| Period | Opioid Deaths (N) | Opioid EMS Calls (N) | Mean Deaths / Week | Mean EMS Calls / Week |
| --- | --- | --- | --- | --- |
| Pre-Intervention (through 2023w25) | 193 | 680 | 1.30 | 4.56 |
| Post-Intervention (2023w26–2025w52) | 74 | 285 | 1.09 | 4.19 |
| **Total** | **267** | **965** | - | - |

The efficacy of GCODE is consistent with recent literature showing that targeted, hot spot-driven naloxone distribution outperforms untargeted community availability in fentanyl-era contexts (Doleac & Mukherjee, 2018; Irvine et al., 2019). The program’s positive findings in the structurally hostile Texas harm reduction environment-characterized by the lowest national OUD treatment density (1.4 per 100,000), syringe service prohibition, and persistent pharmacy-level naloxone barriers (Langabeer et al., 2019; Evoy et al., 2018)-suggest that locally adaptive, digitally coordinated public health action can partially compensate for state-level policy gaps.

An important contextual finding from statewide Texas vital statistics (DSHS, 2025) requires candid discussion. Texas-wide fentanyl poisoning deaths peaked in 2023 (n = 2,307) and declined in 2024 (n = 1,670, −27.6%) and 2025 (n = 1,202, −28.0%), with all unintentional drug poisoning deaths following the same pattern (2023: 5,094; 2025: 3,678, −27.8%). Because this statewide peak and subsequent decline coincides almost precisely with GCODE’s July 2023 deployment, it cannot be ruled out that Galveston County’s improving trajectory partially reflects broader regional or national secular trends rather than the intervention alone. This is a substantive confound that must be stated clearly. At the same time, several considerations attenuate this concern. First, Texas Health Region 6/5S-which includes Galveston County-recorded 526 fentanyl deaths in 2023 (31.5% of the state total), indicating the region bore a disproportionate burden that was not distributed uniformly across the state. Second, the GCODE-attributable slope change in the ITS model is a gradual, sustained weekly trend rather than a simultaneous step-drop, which is more consistent with a cumulative community intervention than a discrete supply-side shock. Third, the statewide decline itself may partially reflect the aggregate effect of local harm reduction programs like GCODE across multiple jurisdictions.

These considerations do not resolve the confounding concern but provide an honest framing: the GCODE findings are best interpreted as consistent with a genuine intervention effect within a broader period of improving opioid mortality, rather than as an isolated county-level success in an otherwise static environment.

The statewide xylazine data add a further layer of supply-side context. Texas documented 42 xylazine-involved poisoning deaths in 2024-nearly triple the 2023 count of 15-before declining to 21 in 2025 (DSHS, 2025). Because naloxone does not reverse xylazine-induced sedation, a local increase in the fentanyl-xylazine combination (“tranq dope”) would reduce the effectiveness of bystander naloxone, while a subsequent decline would have the opposite effect. The timing of this statewide xylazine spike (2024) and decline (2025) introduces an additional unmeasured supply-chain variable that the current ITS model cannot disentangle from the GCODE effect.

Several conditions appear to have enabled GCODE’s effectiveness that may not generalize universally. First, GCODE repurposed digital surveillance infrastructure specifically built for COVID-19 tracking, providing an established data pipeline and interoperability with GAAA that would require significant investment to replicate from scratch. Second, the program benefited from an existing data-sharing agreement between the Health District and GAAA enabling near-real-time EMS linkage. Third, Galveston County’s urban-adjacent density and established network of community organizations provided the organizational substrate needed for rapid naloxone deployment. Fourth, the academic-public health partnership embedded within GCODE provided the analytical capacity for ongoing hot spot identification. These conditions should be explicitly considered when evaluating GCODE’s replicability in rural, lower-infrastructure, or less partnership-dense settings.

Regarding the absence of a concurrent control series: no geographically proximate Texas county with equivalent EMS-to-vital-statistics data linkage was available for comparison, and constructing a synthetic control would require tract-level EMS data that is not publicly accessible. Future studies should prospectively establish matched comparison jurisdictions. Regarding dose–response: granular weekly naloxone dispensation records and active partner site data were not available in the current analytic dataset in a form linkable to the weekly mortality series; this is a critical gap for future GCODE evaluations and a design standard that should be adopted prospectively.

The demographic findings are presented as descriptive. The age distribution of opioid decedents-skewed younger than general mortality-may reflect local epidemiological dynamics specific to Galveston County, though it cannot be rigorously compared to national trends given the data limitations below. Most critically, the severe demographic missingness in the opioid decedent data (41.23% Unknown race/ethnicity) represents a surveillance system failure that precludes meaningful assessment of whether GCODE’s benefits reached communities of color equitably. Because national literature documents pervasive racial disparities in the naloxone care cascade (Khan et al., 2023; Brown et al., 2026), this missingness is not a trivial analytic limitation-it constitutes a fundamental barrier to equity evaluation. Standardizing demographic data collection in overdose surveillance must be treated as a prerequisite for harm reduction equity assessment, not an afterthought.

## Limitations

First, the observational ITS design cannot establish definitive causality. Causal inference is constrained by the potential presence of unmeasured time-varying confounders between 2023 and 2025, including changes in fentanyl supply chain composition and purity, the emergence and subsequent partial retreat of xylazine adulteration in the regional drug supply (documented statewide: 11 deaths in 2021 rising to 42 in 2024 before declining to 21 in 2025; DSHS, 2025), shifts in regional synthetic opioid distribution, concurrent state or federal policy changes affecting naloxone access or OUD treatment availability, and-most importantly-the broader statewide decline in fentanyl-involved mortality that began in 2023 and is documented in Texas vital statistics. The temporal specificity of the slope change consistent with GCODE’s deployment supports a causal interpretation; nevertheless, these possibilities cannot be definitively excluded, particularly in the absence of a concurrent control series.

Second, this analysis did not include a geographic control county or a non-equivalent outcome series such as non-opioid accidental deaths. This design limitation is compounded by a critical contextual finding from Texas-wide vital statistics: statewide fentanyl poisoning deaths peaked in 2023 and declined by approximately 28% per year in both 2024 and 2025 (DSHS, 2025)-a trajectory that nearly coincides with GCODE’s July 2023 deployment and could reflect unmeasured secular trends in the drug supply, national harm reduction policy shifts, or aggregate effects of multiple local interventions operating simultaneously across the state. This co-occurring statewide decline cannot be ruled out as a partial explanation for Galveston County’s improving trajectory, and we note it here with transparency. Future evaluations should prospectively establish a matched comparison jurisdiction with equivalent EMS-to-mortality data linkage to enable formal controlled inference.

Third, the observed fatality ratio (Survival Gap) is an EMS-conditional measure and should be interpreted within that scope. Because successful layperson naloxone reversals may not generate a 911 call, the denominator likely undercounts total overdose events; furthermore, as community naloxone distribution increases, this undercounting may grow, potentially contributing to the observed ratio decline. Table 4 presents absolute counts of deaths and EMS calls by period to support independent evaluation of these trends. The ITS mortality slope change and the Survival Gap both point in the same direction, but as noted in the Discussion, they are subject to shared regional confounders and should be understood as consistent directional evidence rather than independent validation.

Fourth, a dose–response analysis linking naloxone dispensation volume or active partner site counts to weekly mortality outcomes was not feasible with the current dataset. Granular operational metrics were not available in linkable form and individual-site reporting raises de-identification considerations. Future program cycles should prospectively collect these metrics to enable this important test of mechanistic specificity.

Fifth, vital statistics data are subject to the temporal lag and classification limitations of Texas’s decentralized reporting system. This is unlikely to introduce differential measurement error between periods, but may modestly attenuate the precision of effect estimates.

Sixth, demographic missingness in the “Other/Unknown” race/ethnicity category-most likely representing unrecorded or incompletely coded vital statistics entries-precluded a reliable equity analysis of intervention reach across racial and ethnic groups. This is a data infrastructure limitation rather than an analytic one, and its resolution requires standardized demographic data collection at the point of death investigation.

Finally, findings should be generalized with appropriate care. GCODE’s positive results emerged within a specific set of enabling conditions-including repurposed COVID-19 digital infrastructure, an established EMS data-sharing agreement, urban-adjacent population density, and an embedded academic-public health analytical partnership. These conditions may not be readily available in rural or lower-resource settings, and programs seeking to replicate GCODE should plan for meaningful investment in data infrastructure and cross-sector partnership development.

## Conclusion

The findings of this study are consistent with the GCODE intervention having meaningfully reduced opioid-involved mortality in Galveston County, reversing a pre-existing weekly increase of 0.16% to a sustained post-intervention decline of 0.55% per week. The 77.5% relative reduction in the observed fatality ratio among EMS-detected opioid events suggests a decoupling of overdose incidence from fatal outcomes, potentially mediated through increased community naloxone access and bystander activation in targeted high-risk settings. While alternative explanations cannot be definitively excluded and both metrics are subject to shared regional confounders, the consistent directional evidence across the ITS mortality model and the Survival Gap analysis supports the public health value of digitally coordinated, hot spot-driven naloxone distribution. Future evaluations should incorporate concurrent control series, prospective dose–response data on naloxone dispensation, and rigorous demographic data standards to enable equity assessment.

## Data Availability

All data produced in the present study are available upon reasonable request to the authors.

## Notes

**Funding/Conflicts:** The authors declare no conflicts of interest. No external funding was received for this study.

### Competing Interest Statement

The authors have declared no competing interest.

### Funding Statement

This study did not receive any funding.

### Author Declarations

University of Texas Medical Branch Institutional Review Board waived approval for the analysis of secondary public health surveillance data (25-O248).

## References

Ahmed, O., et al. (2025). Mortality due to opioid overdose in the United States: Trends from a CDC WONDER analysis (1999–2024). Population Health Management.

Bernal, J. L., Cummins, S., & Gasparrini, A. (2016). Interrupted time series regression for the evaluation of public health interventions: A tutorial. International Journal of Epidemiology, 45 (3), 887–895.

Bottomley, C., Scott, J. A. G., & Isham, V. (2019). Analysing interrupted time series with a control. Epidemiologic Methods, 8 (1).

Brown, E., et al. (2026). Evaluation of naloxone uptake disparities among harm reduction clients in Rhode Island: A deeper dive using disaggregated race and ethnicity data. Rhode Island Medical Journal.

Claborn, K. R., Samora, J., McCormick, K. A., et al. (2023a). “We do it ourselves”: Strengths and opportunities for improving the practice of harm reduction. Harm Reduction Journal.

Doleac, J. L., & Mukherjee, A. (2018). The moral hazard of lifesaving innovations: Naloxone access, opioid abuse, and crime. Social Science Research Network.

Texas Department of State Health Services (DSHS). (2025). Fentanyl Trends Dashboard: Counts and Percentages of Drug Poisoning-Related Deaths, 2018–2025. Texas DSHS Vital Statistics Section. Retrieved from https://healthdata.dshs.texas.gov/dashboard/drugs-and-alcohol/all-drugs/fentanyl-trends

Evoy, K., Hill, L. G., Groff, L. T., et al. (2018). Naloxone accessibility without a prescriber encounter under standing orders at community pharmacy chains in Texas. JAMA, 320 (18), 1934–1936.

Friedman, J., Mann, N., Hansen, H., et al. (2021). Racial/ethnic, social, and geographic trends in overdose-associated cardiac arrests observed by US emergency medical services during the COVID-19 pandemic. JAMA Psychiatry, 78 (8), 886–895.

Fujita-Imazu, S., Xie, J., Dhungel, B., Wang, X., Wang, Y., Nguyen, P., … & Gilmour, S. (2023). Evolving trends in drug overdose mortality in the USA from 2000 to 2020: an age-period-cohort analysis. EClinicalMedicine, 61.

Ghaddar, T., Ferris, A. H., Mejia, M. C., et al. (2023). Evolving trends in U.S. mortality from opioid overdose: Heroin and beyond. American Journal of Medicine.

Irvine, M., Kuo, M., Buxton, J., et al. (2019). Modelling the combined impact of interventions in averting deaths during a synthetic-opioid overdose epidemic. Addiction, 114 (9), 1601–1613.

Keane, C., Egan, J., & Hawk, M. E. (2018). Effects of naloxone distribution to likely bystanders: Results of an agent-based model. International Journal on Drug Policy, 55, 61–69.

Khan, M. R., Hoff, L., Elliott, L., et al. (2023). Racial/ethnic disparities in opioid overdose prevention: Comparison of the naloxone care cascade in White, Latinx, and Black people who use opioids in New York City. Harm Reduction Journal, 20, 4.

Kiang, M. V., Basu, S., Chen, J., & Alexander, M. J. (2019). Assessment of changes in the geographical distribution of opioid-related mortality across the United States by opioid type, 1999-2016. JAMA network open, 2(2), e190040.

Lalani, K., Bakos-Block, C., Cárdenas-Turanzas, M., et al. (2022). The impact of COVID-19 on opioid-related overdose deaths in Texas. International Journal of Environmental Research and Public Health, 19 (3), 1580.

Langabeer, J., Gourishankar, A., Chambers, K., et al. (2019). Disparities between US opioid overdose deaths and treatment capacity: A geospatial and descriptive analysis. Journal of Addiction Medicine, 13 (1), 70–76.

Salazar, C. I., & Huang, Y. (2021). The burden of opioid-related mortality in Texas, 1999 to 2019. Annals of Epidemiology, 62, 1–9.

Schaffer, A., Dobbins, T., & Pearson, S.-A. (2021). Interrupted time series analysis using autoregressive integrated moving average (ARIMA) models: A guide for evaluating large-scale health interventions. BMC Medical Research Methodology, 21, 58.

Skinner, A., Nolen, S., Cerdá, M., et al. (2024). A simple heuristic for allocating opioid settlement funding to reduce overdose mortality in the United States. The American Journal of Drug and Alcohol Abuse.

Spittal, M., Gunnell, D., Sinyor, M., et al. (2024). Evaluating population-level interventions and exposures for suicide prevention. Crisis.

Walley, A. Y., Xuan, Z., et al. (2013). Opioid overdose rates and implementation of overdose education and nasal naloxone distribution in Massachusetts: Interrupted time series analysis. British Medical Journal, 346, f174.

